# Prenatal diagnosis of sickle cell disease by amniocentesis using FTA technology in a context of precariousness in sub-Saharan Africa: Challenges and perspectives

**DOI:** 10.64898/2026.03.20.26348948

**Authors:** Nelly Ciombo Kamuanya, Vicky Bolamba Lokomba, Emmanuelle Ketsia Bokashanga Mikobi, Helena Trinité Minga Mikobi, Prosper Tshilobo Lukusa, Tite Minga Mikobi

**Affiliations:** Unité de Recherche Clinique Drépanocytose (URC-D), Department of fundamental Sciences, Molecular Biology Service, Faculty of Medicine, University of Kinshasa, DRC; Sickle cell center, IRSS, Kinshasa, RDC; Department of Gynecology and Obstetrics, University Hospital of Kinshasa, DRC; Genetics Unit, Neonatology Service, Department of Pediatrics, University Hospital of Kinshasa, DRC

**Keywords:** prenatal diagnosis, sickle cell disease, amniocentesis, FTA Elute, poverty

## Abstract

Sickle cell disease (SCD) is the most common inherited hemoglobinopathy worldwide. Improving the quality of life of people with SCD requires prenatal and neonatal screening.

Our primary objective was to demonstrate that prenatal diagnosis of SCD is possible even in situations of poverty. Secondarily, we described the socioeconomic profile of couples seeking molecular diagnosis of SCD in Kinshasa, Democratic Republic of Congo.

**Methods:** This was a cross-sectional study conducted in Kinshasa between January 2020 and December 2025. During this study period, 107 couples underwent prenatal diagnosis. Prenatal diagnosis was performed using amniocentesis with FTA Elute technology. This diagnosis was confirmed at birth using cord blood DNA extracted via the conventional salting-out technique.

**Results:** The mean age of the pregnant women was 28 ± 4 years. Eighty-one couples (75.7%) were Christian, nine couples (8.4%) were Muslim, and seventeen couples (15.8%) were animist. Eighty-two couples (76.6%) were known heterozygous AS couples, eleven (10.2%) were heterozygous couples, and fourteen (13.0%) were couples composed of one homozygous SS and one heterozygous AS partner. All pregnancies were singleton. Socioeconomic status was upper middle class (39.2%). The AS genotype was found in 79% of the fetuses. One intrauterine fetal death was observed after amniocentesis. In terms of handling, the FTA Elute technology reduces DNA extraction time to 30 minutes. It is easy to use. Results are available in less than 24 hours.

**Conclusion:** The FTA Elute technology is a reliable, less expensive, and easy-to-use prenatal screening technique for sickle cell disease. Sample transport and storage conditions are better suited to resource-limited settings.

## Introduction

It is the most widespread hemoglobinopathy in the world and in sub-Saharan Africa in particular [1 - 3]. The World Health Organization (WHO) estimates that there are approximately 350,000 new births of sickle cell patients worldwide each year. Of these new births, 80% occur in sub-Saharan Africa [2, 3]. In the Democratic Republic of Congo (DRC), the prevalence of sickle cell trait carriers is estimated at between 25 and 35%, while homozygous SS individuals represent 1.7 to 2% of the population. The DRC is the second most affected african country by SCD after Nigeria. Clinically, SCD is associated with high early mortality. Acute and chronic complications are associated with very high morbidity. Indeed, in sub-Saharan Africa, this early mortality reaches between 40 and 55% before the age of five [4-6]. There is no etiological treatment for SCD. Bone marrow transplantation and hematopoietic stem cell therapy are currently available as replacement therapies capable of permanently curing the symptoms of sickle cell disease. However, they cannot be used as a mass treatment. These treatments are neither accessible nor available in resource-limited countries. Hydroxyurea is the only drug that has shown efficacy in improving the quality of life of patients [7–9]. Once again, hydroxyurea is not yet accessible and available in many resource-limited african countries, including the DRC [9, 10]. The best management of SCD begins with early diagnosis. Indeed, early detection allows for the prevention of acute complications in the short term and a reduction in the frequency of debilitating chronic complications in the long term. Early diagnosis can be neonatal or prenatal. Neonatal diagnosis uses several techniques, including molecular testing, but prenatal diagnosis relies exclusively on molecular testing, which is the only confirmatory test. Amniocentesis allows for the collection of amniotic fluid from which fetal cells can be obtained, from which fetal DNA can be extracted and analyzed. Typically, DNA from fetal cells is extracted using conventional techniques, including the salting-out technique described by Miller et al. [11]. Currently, several commercial DNA extraction kits are available. These techniques require a DNA extraction step lasting between 6 and 8 hours, followed by a DNA amplification step (PCR step) and finally a post-PCR step. These conventional techniques require specific equipment at each diagnostic stage, and the turnaround time for results is 48 hours. Currently, new technologies make it possible to reduce not only the number of steps in the molecular diagnosis process of sickle cell disease, but also the cost of the test and the time to deliver the result are substantially reduced. The objective of this study was to demonstrate that prenatal diagnosis of SCD is possible in a resource-limited setting using less expensive techniques. Secondarily, we aimed to determine the socioeconomic profile of couples requesting molecular diagnosis of sickle cell disease in Kinshasa, DRC.

## MATERIALS AND METHODS

We conducted a cross-sectional study between december 2020 and december 2025. The study was conducted in two hospitals in Kinshasa: the sickle cell center, a public center specializing in the management of SCD in Kinshasa, and the university hospital of the School of Medicine of the University of Kinshasa. During the study period, we received 107 couples requesting prenatal diagnosis of SCD. Eighty-two couples were referred to us by their primary care physician, and twenty-five couples received genetic counseling. All couples had at least one child with SCD.

### Inclusion criteria

In this study, we included pregnant women from couples requesting prenatal diagnosis, referred by a treating physician, or who received genetic counseling during the study period. All pregnant women had signed an informed consent form.

### Operational Definitions

In our study, the households of pregnant women were classified into five socioeconomic classes according to the criteria we defined below. These included elite households, upper-middle-class households, lower-middle-class households, working-class households, and low-income households. The operational definitions of these different socioeconomic classes are given below.

### Elite class households

households with very high wealth and income. These include top business executives, senior civil servants, high-profile professionals, or heirs.

### Upper middle class households

these are households composed of managers and higher intellectual professions, such as engineers, doctors, lawyers, executive managers, university professors.

Lower middle class households: these households include intermediate professions such as secondary school teachers, nurses, technicians, supervisors, small business owners.

Working-class households include households with more modest incomes, often linked to manual or operational work such as skilled or unskilled workers, service employees (cashiers, maintenance workers), and drivers.

Households in the precarious class include households living below the poverty line or dependent on social assistance such as the long-term unemployed, precarious workers, and the homeless. Amniotic fluid sampling and sickle cell disease diagnostic technique

The amniotic fluid sample (15 ml) was collected by amniocentesis between the 16th and 17th weeks of amenorrhea. The molecular diagnosis of sickle cell disease was made using the FTA Elute technology.

The FTA Elute cards were kindly provided by the Center for Human Genetics at Gasthuisberg University Hospital of the Catholic University of Louvain (Belgium). Molecular analyses were performed at the laboratory of the Center for Human Genetics of the School of Medicine, University of Kinshasa. A second molecular test was performed at delivery using the newborn’s umbilical cord blood and the classic salting-out technique described by Miller et al. [11].

### FTA Elute Technology

FTA Elute is a card impregnated with a patented chemical process. When a biological sample is placed on the FTA Elute card, this chemical process lyses the cells and denatures the proteins, while the nucleic acids are extracted and protected within the paper fibers. The FTA Elute card can then be transported at room temperature to the laboratory or stored at room temperature for later analysis. DNA is recovered in solution after a simple 30-minute water elution step, while PCR inhibitors are retained within the FTA Elute board matrix. Following this elution step, the manufacturer directly performs the PCR reaction. FTA Elute is a true extraction kit, enabling sample collection, transport, and room-temperature DNA storage with reduced transportation costs. FTA Elute technology eliminates the risk of contamination for those handling the sample. As a result, the sample is no longer classified as “biohazardous” and can be mailed. FTA Elute technology eliminates lengthy and time-consuming extraction procedures, ranging from 4 to 16 hours, and reduces the cost of reagents for the DNA extraction step. For hemoglobin PCR analysis, FTA Elute delivers a DNA solution free of PCR inhibitors. Laboratory Processing of the Amniotic Fluid Sample

After amniocentesis, the amniotic fluid sample was taken directly to the laboratory for analysis. In the laboratory, the tube containing 15 ml of amniotic fluid was labeled with the data obtained from the pregnant woman. The sample was then centrifuged at 3500 rpm for 30 minutes. After centrifugation, the supernatant was discarded. We added 100 µl of PCR water to the amniotic fluid sediment; then we homogenized the sediment with the 100 µl of PCR water. Using a micropipette, we aspirated 40 µl of the mixture and spotted it onto the FTA ELUTE card at four different locations specified by the manufacturer. We dried the FTA ELUTE card at room temperature for one hour.

### Sample Analysis Technique on FTA Elute

Using a suitable punch, we cut 1.2 mm from the FTA Elute card at the area covered by the dried amniotic fluid. The cut punch was placed in a PCR tube. After punch collection, we decontaminated the punch by cutting a white punch from an area of the FTA Elute card not covered by the amniotic fluid sample. The first step of the analysis begins with the elution or washing of the sample punch by adding 500 µl of PCR water to the PCR tube. We then vortexed the tube three times for 5 seconds each time. Using a sterile micropipette tip, we removed the punch by pressing it against the wall of the PCR tube to remove excess liquid. The liquid sample in the PCR tube is now ready for amplification.

### Sickle Cell PCR

The molecular diagnostic technique for sickle cell disease that we used is based on the principle of detecting the E7V (rs334) sickle cell mutation on the mutated HBB gene. We used the PCR-RFLP technique. We amplified a 440 bp fragment of the HBB gene using the following primers: Forward: - TGTGGAGCCACACCCTAGGGTTG-, Reverse: -CATCAGGAGTGGACAGATCC-. These primers were provided by Integrated DNA Technologies. Amplification of the fragment was performed using an ABI Veriti 96 well thermocycler. The amplified fragment of interest contains two *DdeI* restriction sites. In the case of the E7V mutation, the first DdeI restriction site is eliminated. Diagram 1 below shows the amplified 400 bp fragment of the HBB gene and its DdeI restriction sites.

**Diagram-1 :**
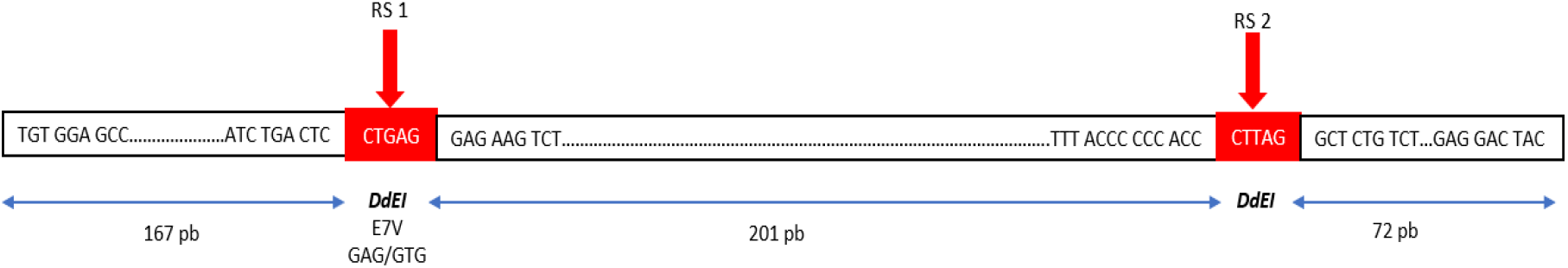
Fragment of interest of the HBB gene in sickle cell disease. **Legend:** RS: restriction site

### Amplification Program

Our amplification program consists of three traditional PCR steps: initiation or denaturation at 95°C for 5 minutes, hybridization at 58°C for 30 seconds, and elongation (extension) at 72°C for 5 minutes. The total number of cycles was 32.

Enzymatic digestion and visualization of the enzymatic digestion product using a UV transilluminator After amplification of the HBB gene fragment of interest, we proceed to the enzymatic digestion stage. The restriction enzyme used was *DdEI*. In the absence of the E7V mutation (homozygous AA subject), the DdEI enzyme recognizes both restriction sites. Thus, after PCR migration on agarose gel, a homozygous subject (AA) has three bands of the amplified fragment. These three fragments are 72 bp, 167 bp, and 201 bp, respectively. In the presence of the E7V mutation on a parental allele (heterozygous subject (AS)), the DdEI enzyme will recognize one restriction site on the fragment carrying the mutation and two restriction sites on the fragment not carrying the mutation. Thus, after PCR migration on agarose gel, the heterozygous subject AS will have four bands of the amplified fragments. These bands have 72 bp, 167 bp, 201 bp, and 368 bp respectively. Finally, in the presence of two E7V mutations on the two parental alleles, the restriction enzyme will recognize only one restriction site on each fragment carrying the mutation. After agarose gel electrophoresis, the homozygous individual (SS) has two bands on the amplified fragments. These bands have 72 bp and 368 bp respectively.

### Interpretation of results

Our agarose gel was concentrated at 2%. A sample with two bands of 72 bp and 368 bp of the HBB gene on the migration gel was diagnosed homozygous SS, the sample with three bands of 72 bp, 167 bp and 201 bp of the HBB gene was diagnosed homozygous AA, finally the sample with four bands of 72 bp, 167 bp, 201 bp and 368 bp of the HBB gene on the migration gel was diagnosed heterozygous AS.

### Postnatal genotype confirmation

After birth, newborn genotypes were checked using another molecular test involving umbilical cord blood. The technique used for newborn genotype checking was salting out, as described by Miller et al. [7]. This technique extracts fetal DNA from cord blood and requires 6 to 8 hours for extraction before the PCR step can begin. After the DNA extraction stage, the following steps are performed: PCR (lasting 3 to 4 hours), enzymatic digestion (also lasting 3 to 4 hours), and finally, the migration of the PCR product on an agarose gel (lasting 2 to 3 hours).

#### Statistics

Our data was entered into Excel. We determined the mean and standard deviation of the pregnant women’s ages. We also calculated the genotype frequencies of the newborns. Our graphs were created using Excel.

## RESULTS

1. Socioeconomic and demographic characteristics of couples requesting prenatal diagnosis of SCD

The mean age of the pregnant women was 28 ± 4 years, and the mean parity was 2 ± 1.2. The mean age of the husbands was 36 ± 2 years. The mean number of children with known sickle cell disease per couple was 2. Eighty-one couples (75.7%) were Christian, nine couples (8.4%) were Muslim, and seventeen couples (15.8%) had no religion or practiced animism. Eighty-two couples (76.6%) were heterozygous AS couples who knew their hemoglobin genotype during premarital examinations, eleven (10.2%) were heterozygous couples discovered after marriage at the birth of the first sickle cell child, fourteen (13.0%) were couples composed of one homozygous SS and one heterozygous AS. All pregnancies were singleton.

Figure-1 above shows the distribution of the socioeconomic level of households in our study. It appears that the majority of households were distributed between the upper middle class (39.2%), the lower middle class (36.4%), and the elite class (5.6%).

**Fig 1:**
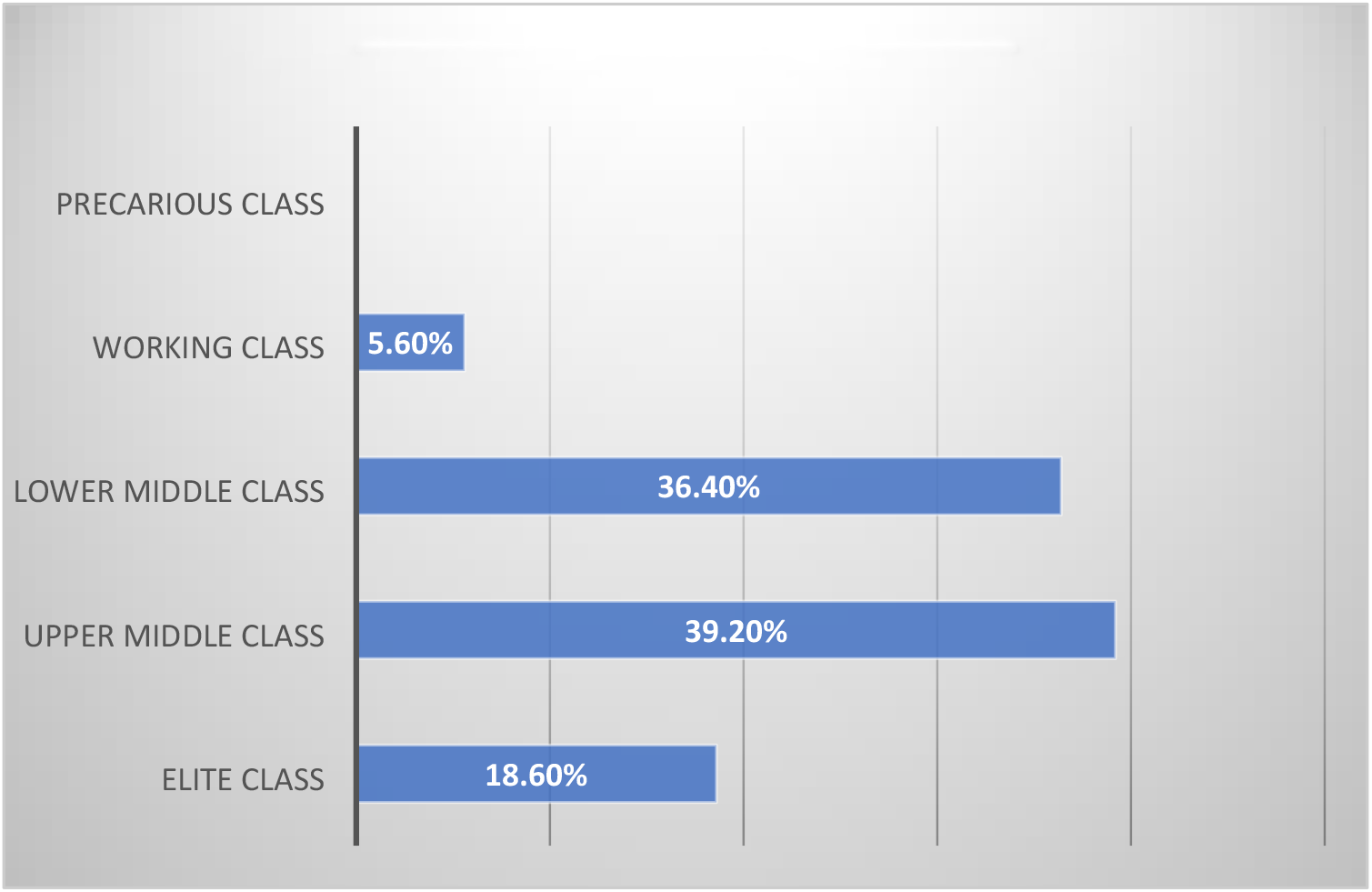
distribution of the socioeconomi level of families

3. Genotype frequencies after molecular testing by amniocentesis

Figure 2 above shows the frequency of fetal genotypes after molecular testing by amniocentesis in our study. We note that 77% (82 fetuses) had the AS genotype, 14% (15 fetuses) had the AA genotype, and 9% (10 fetuses) had the SS genotype.

**Fig 2.**
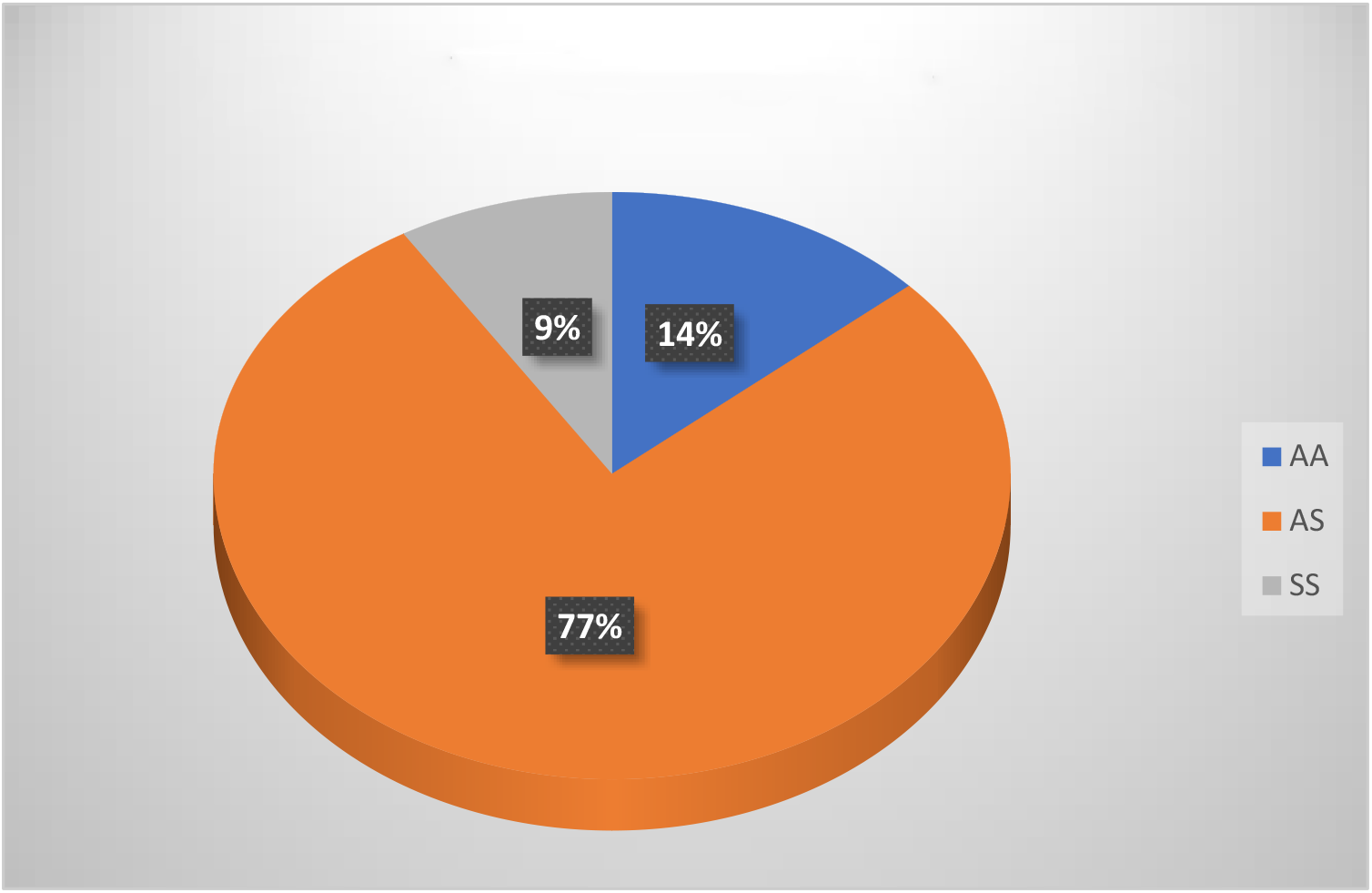
Genotype frequencies after molecular testing by amniocentesis

Figure 3 above shows the different genotypes after diagnosis using FTA Elute technology (gel on the left) and the salting-out technique from umbilical cord blood (gel on the right). Moving from left to right in Figure 3, we have the column of molecular weight markers (MWM), followed by the negative control: water (NCH2O). We then have three positive controls: homozygous AA (PC1), heterozygous AS (PC2), and homozygous SS (PC3). Finally, the amniocentesis samples showed three genotypes similar to the control genotypes. These were the genotype of sample SP1, corresponding to a fetus with genotype AA; sample SP2, corresponding to a fetus with genotype AS; and sample SP3, corresponding to a fetus with genotype SS. These results were confirmed with umbilical cord blood at birth using the salting-out technique (gel on the right).

**Fig 3:**
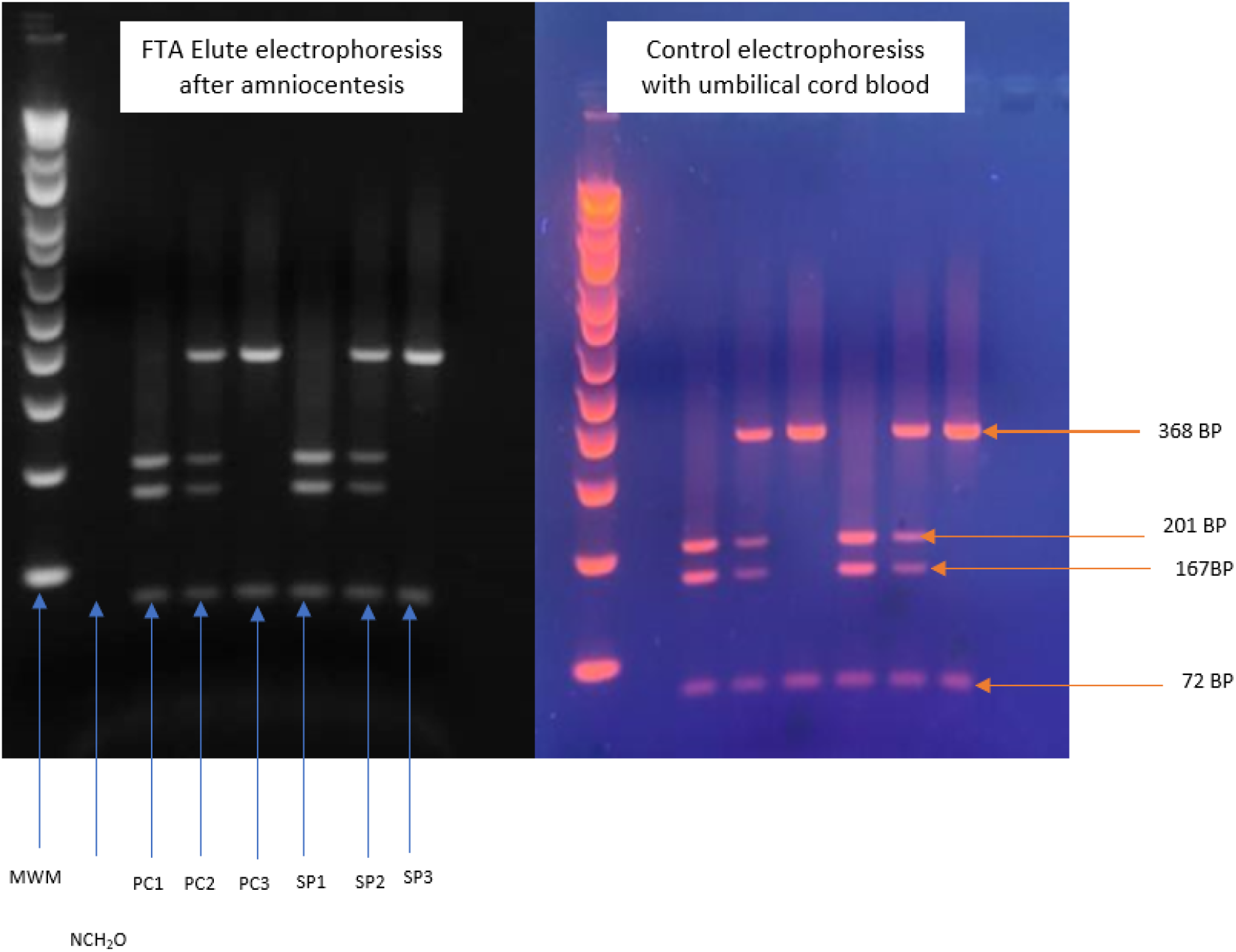
DNA electrophoresis profile on agarose gel using FTA Elute technology and the conventional salting-out technique. **Legend** : ***MWW***: molecular weight markers; ***NCH2O***: H2O negative control; ***PC1***: AA homozygous positive control; ***PC2***: AS heterozygous positive control; ***PC3***: AA homozygous positive control; ***SP1***: AA homozygous positive sample; ***SP2***: AS heterozygous positive sample; ***SP3***: SS homozygous positive sample.

Figure-4 : above shows the distribution of incidents during pregnancy after amniocentesis. We observe that 99% (106 pregnancies) resulted in live births, while 1% (one pregnancy) resulted in a stillbirth.

**Fig 4:**
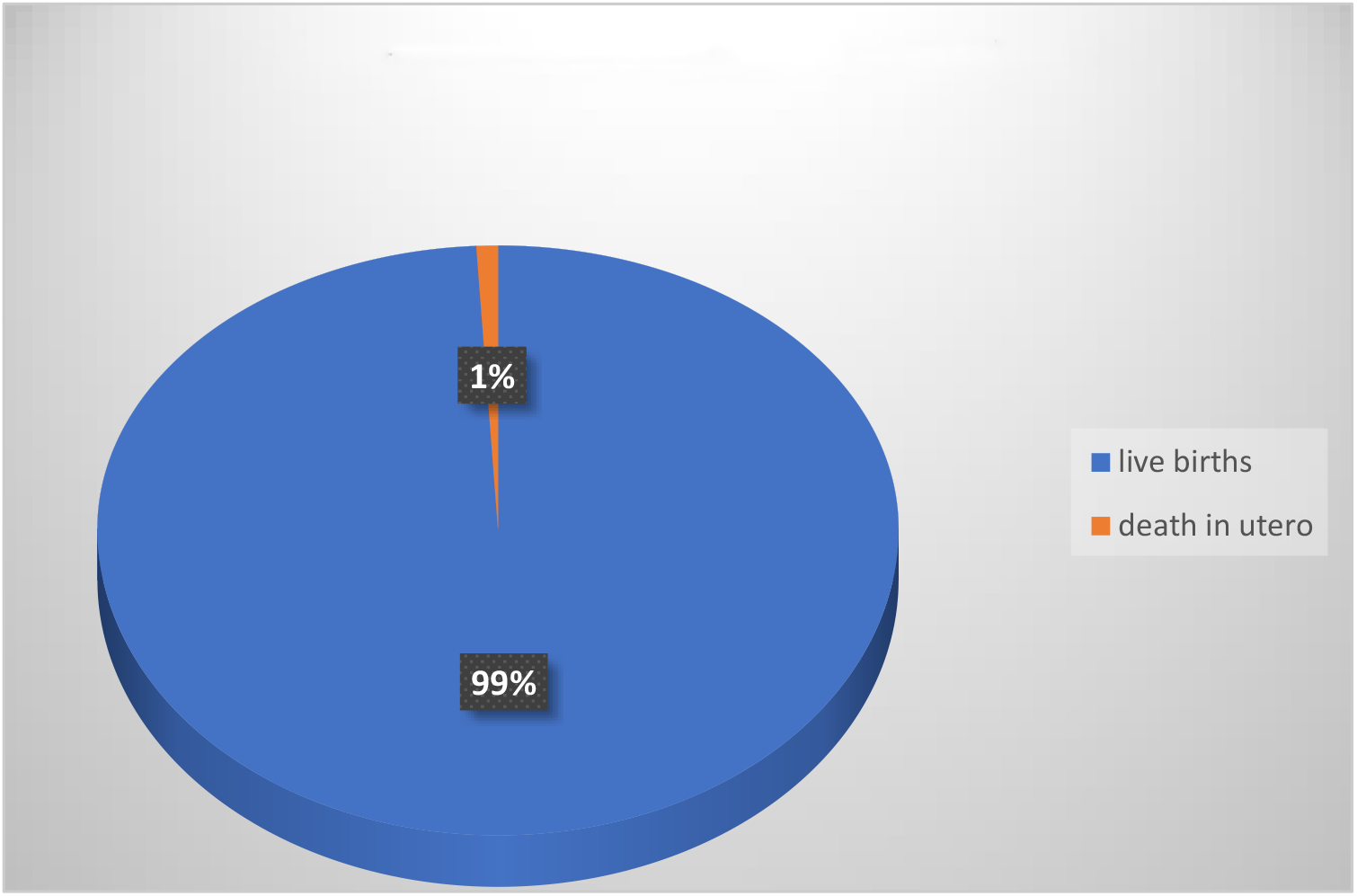
Pregnancy outcomes after amniocentesis**99%**

Figure-5 above shows the cost-specificity ratio between FTA elute technology and commercial kits. We observe that, for the same specificity, the FTA elute card used in nucleic acid extraction is less expensive than the reagents used in commercial kits; it is easier to manipulate the DNA extraction steps with FTA elute technology than with commercial kits; the DNA extraction time is reduced from 8 hours to 30 minutes; the cold chain for sample storage and transport is eliminated; and the results are available in less than 24 hours compared to other analytical techniques.

**Fig 5:**
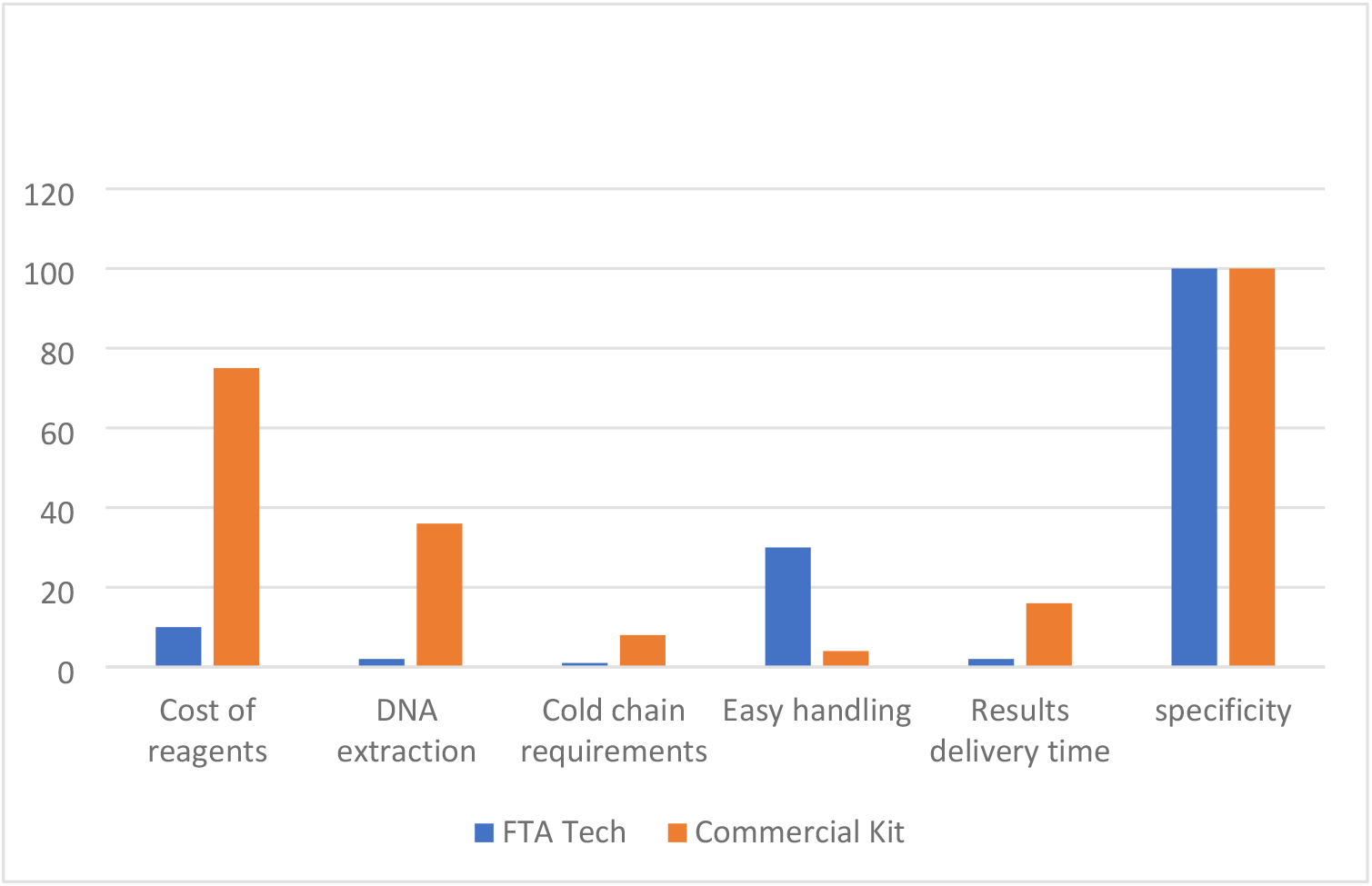
Cost-effectiveness ratio of the FTA Elute technology and commercial kits

## DISCUSSION

Early screening for sickle cell disease is a strategy that has contributed to improving the early management of sickle cell patients. Early management has led to improved quality of life for patients and a reduction in morbidity and mortality. Our study aimed to demonstrate that prenatal diagnosis of sickle cell disease is possible in a context of poverty using less expensive techniques.

Our study showed that couples requesting prenatal diagnosis of sickle cell disease are young, with an average age of 28. This age is close to that found in other African studies for prenatal screening of hemoglobinopathies: 32 years in Tunisia [12], 31 years in Northern Cyprus [13]. Religious affiliation does not influence the use of prenatal screening for sickle cell disease. In this study, the predominance of the Christian religion may be due to the fact that the DRC is a predominantly Christian country. Knowledge of the hemoglobin genotype does not influence marriage. Indeed, 76% of couples in our study were informed of their hemoglobin genotype during premarital examinations. This late knowledge of the hemoglobin genotype justified the use of prenatal screening for sickle cell disease. In our series, 10% of couples were informed of their hemoglobin genotype after the birth of a child with sickle cell disease. Thus, to prevent the birth of another child with sickle cell disease, they resorted to prenatal diagnosis. Our study also showed that knowledge of hemoglobin genotype does not influence marriage decisions. In fact, fourteen couples consisted of one partner with a known homozygous SS genotype and the other with a known heterozygous AS genotype. We believe that a lack of information and socioeconomic status are responsible for these high-risk types of marriages. Indeed, marriages between a homozygous SS partner and a heterozygous partner were observed in working-class couples, which include skilled and unskilled laborers and service workers.

From a socioeconomic and professional perspective, our study showed that the majority of couples requesting prenatal diagnosis of sickle cell disease belonged to the upper or lower middle class and the elite. This observation may be due to the fact that these two social classes are the best informed about the disease and can afford early diagnosis, particularly prenatal diagnosis.

In terms of hemoglobin genotype frequencies, our study identified 10 fetuses (9%) with the SS genotype. This frequency clearly demonstrates the risk that a heterozygous AS couple has of having a homozygous SS child.

Regarding pregnancy outcomes after amniocentesis, we observed one stillbirth (1%). Amniocentesis is a safe procedure, but it does carry some complications, including miscarriage (0.5 to 1%), the risk of infection, and premature rupture of membranes [15]. The risk of miscarriage is very low [14, 16]. In our series, the stillbirth occurred in a pregnancy of 33 weeks of gestation. This incident was not directly related to the amniocentesis procedure, as it occurred in the context of two maternal episodes of malaria with high parasitemia and a maternal urinary tract infection with multidrug-resistant organisms.

In our study, the cost-specificity ratio of FTA Elute technology and other commercial kits showed that both techniques have 100% diagnostic specificity. However, FTA Elute technology offers many advantages for users in resource-limited countries. Indeed, the FTA Elute card is less expensive, and the technology eliminates the DNA extraction step, which is very time-consuming (approximately eight hours with other techniques) and requires very expensive equipment. FTA Elute technology eliminates the need for a cold chain during sample storage and transfer. FTA Elute technology is easy to use and reduces the turnaround time for results by 24 hours.

## CONCLUSION

In high-resource countries, screening for congenital and genetic diseases that lead to disability and early death in newborns is recommended. Prenatal diagnosis is the only way to prevent the birth of children with congenital and genetic diseases. The FTA Elute technology offers a reliable, less expensive, and easy-to-use prenatal screening technique for sickle cell disease. It should be widely adopted in resource-limited countries with a high prevalence of sickle cell disease. Resource-limited countries must promote prenatal screening programs to reduce the prevalence of sickle cell disease and ensure public education and awareness. FTA Elute technology offers optimal conditions for sample transport and preservation through simple air drying and storage under the same conditions, eliminating the need for a cold chain. This minimizes turnaround time for results. With the same specificity as other prenatal sickle cell screening techniques, FTA Elute technology provides resource-limited countries with significant savings in both cost and time.

## Data availability

All data relating to this manuscript are available upon request from the authors..

## Ethical approval

The study was approved by the ethical committee of the school of public health of the University of Kinshasa (Approval reference: FACMED/UNIKIN/CE/015/2020), DRC. Informed consent was obtained from all patients before their inclusion in the study.

## Acknowledgments

the authors thank all the pregnant women who consented to participate in this study, as well as the laboratory technicians who analyzed the samples. We also thank the Human Genetics Laboratory at KU Leuven for generously providing us with the FTA Elute cards and the nurses who provided social support to all the pregnant women during the examination period.

## Authors’ contribution

**NCK**: data collection, sample analysis, interpretation of results, genetic consultations; **VBL**: genetic consultations, manuscript editing, amniotic fluid sampling, **EKBM** : data collection, genetic counseling; **HTMM**: data collection, genetic counseling; **PTL**: genetic counseling, manuscript editing, **TMM:** topic design, manuscript writing, amniotic fluid sampling.

## THE BULLETED STATEMENT

### What’s already known about this topic?

Prenatal diagnosis is used to screen for diseases with high morbidity and mortality. Conventional techniques using commercial kits are employed. The use of FTA elute technology has not yet been reported for this type of diagnosis in resource-limited settings.

### What does this study add?

This study is the first in sub-Saharan Africa to use FTA elute technology in the prenatal diagnosis of sickle cell disease, and it proves that this diagnosis is possible in resource-limited countries.

